# Increasing family participation in physiotherapy-related tasks of critically ill patients: a pilot study on the feasibility and preliminary effectiveness of an intervention

**DOI:** 10.1101/2025.10.27.25338462

**Authors:** Lotte (LMM) van Delft, Karin (K) Valkenet, Arjen (AJC) Slooter, Cindy (C) Veenhof

**Affiliations:** Department of Rehabilitation, Physiotherapy Science and Sport, University Medical Center Utrecht, Utrecht University, Utrecht, The Netherlands; Research Group Innovation of Human Movement Care, HU University of Applied Sciences Utrecht, Utrecht, The Netherlands; Department of Intensive Care Medicine, University Medical Center Utrecht, Utrecht University, Utrecht, The Netherlands

**Author notes:** Corresponding author: Karin Valkenet.

## Abstract

**Objective:** To investigate the feasibility and preliminary effectiveness of an intervention for family participation in physiotherapy-related tasks of critically ill patients.

**Design:** This quasi-experimental pilot study with a non-equivalent control group was conducted within a mixed adult Intensive Care Unit (ICU). Included were first-degree relatives of critically ill patients who were admitted to the ICU for more than three days and received physiotherapy.

**Study parameters:** The primary outcome was feasibility, based on a minimal 1) recruitment rate; 2) percentage of relatives who chose physiotherapy-related tasks; 3) usage of physiotherapy-related tasks; 4) usability; and 5) maximum number of adverse events. Secondary outcomes were patients’ physical functioning and symptoms of anxiety, depression and post-traumatic stress disorder in family members, at ICU discharge.

**Results:** Between March and June 2021, 34 family members (19 intervention, 15 control) of 27 critically ill patients (16 intervention, 11 control) were included. The recruitment rate was 75.6% (34 out of 45). The percentage of family members who chose physiotherapy-related tasks at baseline was 94.7% (18 out of 19). The average percentage of days that these family members performed physiotherapy-related tasks was 56.7%. The mean System Usability Scale score was 77.1. No adverse events occurred. At ICU discharge there was a significant difference in the mean Hospital Anxiety and Depression Scale - depression score (P=0.030) and mean Impact of Event Scale – Revised (P=0.047) score between the intervention and control group, in favor of the intervention group.

**Conclusion:** Family participation in physiotherapy-related tasks of critically ill patients seems feasible.

## 1. Introduction

Admission to an Intensive Care Unit (ICU) is often accompanied by the emergence of ICU acquired weakness [1]. One year after discharge, more than 50% of the patients treated for more than two days in the ICU still have physical limitations in their daily functioning [2-4]. Additionally, survivors of critical illness often experience long-term impairments in cognitive function and/or mental health [4-7]. In 2012, the Society of Critical Care Medicine introduced the term “Post-Intensive Care Syndrome” (PICS) to describe these impairments arising after critical illness [7]. The term PICS can be applied to an ICU survivor but also to a family member of a patient who had been admitted to an ICU (PICS-F), since the psychological impact of an ICU admission is not limited to the patients, but appears to affect the mental health of relatives as well [7]. Family members are at high risk of anxiety, depression, a posttraumatic stress disorder (PTSD), and complicated grief, which adversely effects quality of life of the whole family [8-10].

A crucial element of daily ICU care is early mobilization and exercise [11-13]. The last 20 years evidence confirmed the benefits of ICU early rehabilitation, which can improve patients’ physical function, shorten length of stay, and reduce adverse psychological effects [11, 13-15]. Another important component in ICU care is family engagement; family engagement can be beneficial for both patients and their family members [16, 17]. Engaging families could humanize ICU care, may enhance psychological wellbeing for both patients and family and improve family members’ ability to cope with the patients’ situation by having a purposeful role [16, 18]. Since physical therapists treat patients mostly once a day and nurses often lack time to help patients with their exercises, active family involvement may support staff by utilizing family members as a supporting resource to deliver additional care [17]. Currently, family involvement in the ICU mostly involves improving communication and dissemination of information, or participation in nursing activities. However, family members may also be engaged in many practical low-cost, high-value rehabilitative physical activities of critically ill patients [16-20]. At this moment, limited research has been performed on the feasibility and benefits of family participation in physiotherapy-related tasks, and there are currently no interventions developed on this topic [20].

For that reason, a multidisciplinary team of experts developed a new intervention, aiming to increase family participation in physiotherapy-related care of critically ill patients with the goal of improving patients’ physical functioning and reducing stress-related symptoms in family members. For the development of this intervention, the Medical Research Council (MRC) framework for developing and evaluating complex interventions was used [21]. After designing a new complex intervention, a feasibility study is necessary before conducting large scale evaluations because these are often undermined by problems of feasibility, usability, and recruitment [21, 22].Therefore, the primary aim of this study was to investigate the feasibility of family participation in physiotherapy-related tasks of critically ill patients. Secondly, this study aimed to evaluate preliminary effectiveness of the intervention on patients’ physical functioning and symptoms of anxiety, depression and PTSD in family members.

## 2. Methods

### 2.1 Design, setting and sample

This was a quasi-experimental pilot study with a non-equivalent control group design, conducted within a mixed medical-surgical-cardio-neuro-adult ICU of the University Medical Center Utrecht, the Netherlands. During this pilot, up to 16 of the 32 admission spaces were occupied by isolated COVID-19 patients, and the opportunities for family members to visit were limited to a maximum of two different people per 24 hours. Participants of this study were first-degree relatives or spouses of critically ill patients who were admitted in the ICU for more than three days and received physiotherapy as daily care. In order to be eligible to participate in this study, both the family member and the patient had to meet all inclusion criteria, and none of the criteria for exclusion (table 1). Per patient, a maximum of two family members were allowed to participate in this study. A sample size of at least 30 family members (15 in both the intervention and control group) was determined in accordance with published guidance for pilot studies [22]. Participants were given information about the intervention and its aim, followed with a choice whether they wanted the intervention or not. If not, they entered the control group. Once the sample of one of the two groups was reached, the inclusion proceeded to reach the sample of the other group only. The study protocol (METC number 21-071) was approved by the Medical Ethics Committee of the University Medical Center Utrecht. All family members provided written consent for their involvement in this study. Consent was not required from patients because family members were asked to sign a broad consent to use patient data collected in usual care for medical research.

**Table 1.** Inclusion and exclusion criteria for participation in this pilot study.

|  | Inclusion criteria | Exclusion criteria |
| --- | --- | --- |
| **Patients** | - ICU admission for at least three days<br>- receiving physiotherapy: ICU patients receive physiotherapy from day three. Sedated patients two times a week and awake patients 3-5 times a week<br>- expected to remain in the ICU for more than two days | - aged < 18<br>- COVID-19 (isolation) patients<br>- patients with increased intracranial pressure<br>- receiving end of life care<br>- no family/contact person |
| **Family** | - first-degree relative, spouse or other first contact person<br>- able to speak and read Dutch |  |

### 2.2 Intervention development

An intervention was developed by a multidisciplinary team of experts, using an evidence and expert based, iterative process following the MRC framework [21]. The first step of the development was to identify existing evidence by executing a mixed-methods systematic review [20]. The following step was identifying and developing theory, by performing a qualitative study, to gain knowledge on the area of concern, to get an understanding of the future users and their context, and determine which values and requirements the different stakeholders deem important to include in the intervention. Therefore, a qualitative study, using semi-structured interviews with critically ill patients, their family and staff members, was conducted. The interviews demonstrated that patients, their family and staff members have a positive attitude towards the participation of family members in physiotherapy-related care of critically ill patients [19]. Additionally, important components and requirements for the future intervention were collected [19]. As last step in the development phase, all ideas and requirements were merged and prototypes were developed by the project team. Several questionnaires were administered to test these prototypes and procedures with nurses and physiotherapists, as well as critically ill patients and their loved ones. After the prototyping phase, the intervention for this pilot study was developed and validated by the project team, the physiotherapists working in the ICU and some family members.

### 2.3 The intervention

The final intervention consists of three components. The first component is a brochure, including information about the possible physical and psychological impact of an ICU admission on both critically ill patients and their loved ones, and the concept of family participation in physiotherapy-related tasks. In addition, this brochure contains a menu with activities for family members to participate in (table 2). The activities are divided in category A and B. Category A are simple activities that family members can do to calm the patient (e.g. reading a book, or massaging the hands/feet). Category B are physiotherapy-related tasks including: passive movement exercises (range of motion), active limb exercises, functional exercises and breathing exercises. All activities in the menu are optional, therefore the menu also contains the option “I do not want to participate (yet)”. In the brochure, the physiotherapy-related tasks are written out in more detail and supplemented with QR codes, linked to videos in which the physical exercises are demonstrated. As second component of the intervention, a poster for the patients’ room was developed. On this poster all possible activities for family members to participate in, were listed. In the first column the preferred activities are noted by the family members. Thereafter, the physiotherapist marks, after each physiotherapy treatment, exercises appropriate for the patients’ functioning in the second column. Subsequently, during the admission, family records when and how often they performed which activities in the third column. This poster makes it visible to everyone in which activities family wants to participate, which of these activities are deemed suitable for family participation by the physiotherapist, and what the frequency of performed activities is. An example of the poster (in Dutch) can be requested via the corresponding author. The last component of the intervention is a real life personalized instruction moment with a physiotherapist. During this meeting the activities are explained and if necessary practiced together.

**Table 2.**
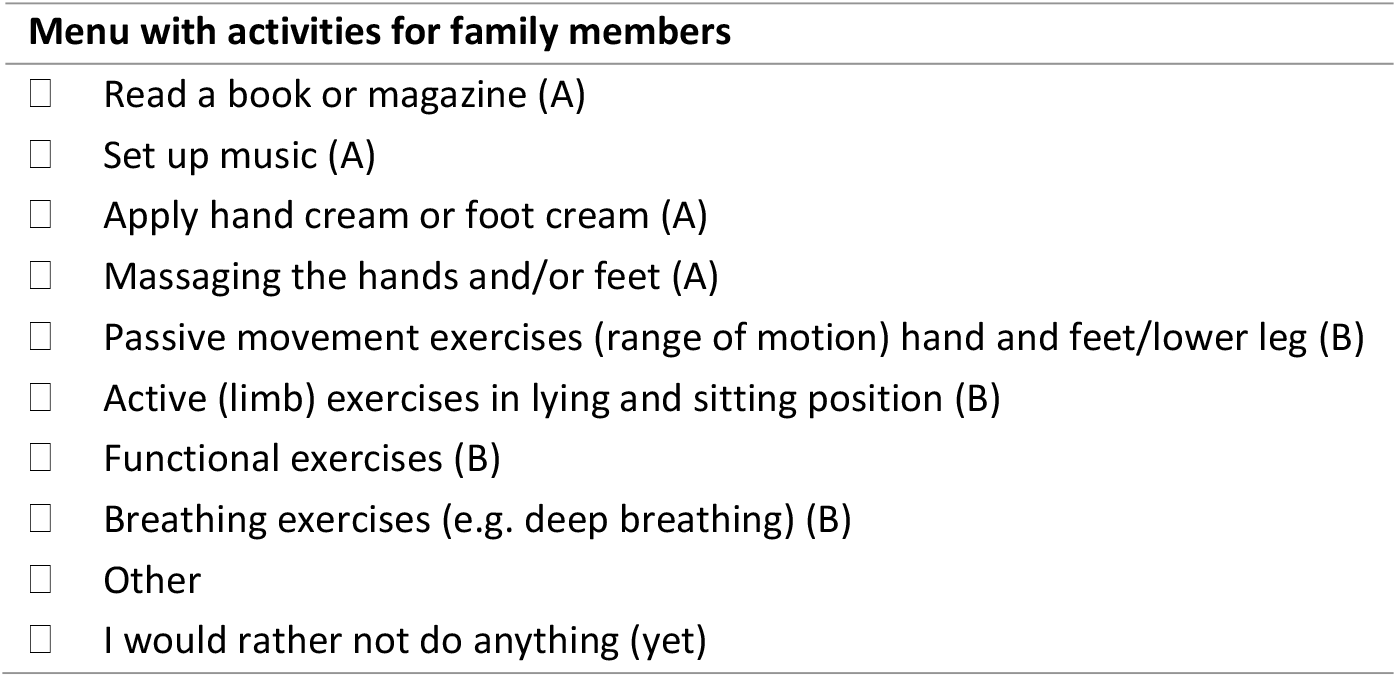
Menu with activities divided in category A (activities to calm the patient) and B (physiotherapy-related tasks)

### 2.4 Study procedure and data collection

After the first physiotherapy consult, standard at day three or four of ICU admission, the physiotherapist checked whether the patient met the inclusion criteria, and asked the nurse in charge whether there were family members who met the inclusion criteria. If yes, the main investigator (LvD) approached them (real life or by phone) to provide information about the study, and asked for informed consent. If family wanted to participate, family members received the brochure and an appointment with the physiotherapist for instructions in the upcoming two days after inclusion. During that meeting, the brochure and poster were explained, activities were chosen and if necessary exercises were practiced together. After each physiotherapy treatment, the physiotherapist marked the suitable exercises for family members to perform on the poster. Family members were asked to report the executed activities on this poster by recording when and how often they performed which activities, and write down if something unintended happened with medical devices (e.g. dislocation of endotracheal tube or lines) or with the patient (e.g. injuries). Three days after starting, and thereafter once a week, the main investigator evaluated with the family member (phone or real life) if there were any questions or issues to discuss.

#### 2.4.1. Data collection in both groups

As shown in figure 1, demographics of patients and family members were collected as part of usual care. Collected characteristics of family members were: age, sex, relation to the patient and whether or not there was a previous experience in the ICU (i.e. admission or work-related). Collected characteristics of patients were: age, sex, date of ICU admission, type of ICU admission (i.e. planned surgery, medical, emergency surgery), Acute Physiology And Chronic Health Evaluation (APACHE) II score, highest Sequential Organ Failure Assessment (SOFA) score, length of stay and discharge location. At discharge, patients’ physical functioning was reported by the physiotherapists, as part of usual care, using the MRC-sum score and ICU Mobility Scale (IMS) [1, 23]. In addition to these data from usual care, at ICU discharge, symptoms of anxiety and depression in family members were measured with the Dutch version of the Hospital Anxiety and Depression Scale (HADS) [24, 25] and PTSD-related symptoms were measured with the Dutch version of the Impact of Event Scale – Revised (IES-R) [26-28]. These two surveys were sent to family members by email, to be completed privately.

**Figure 1.**
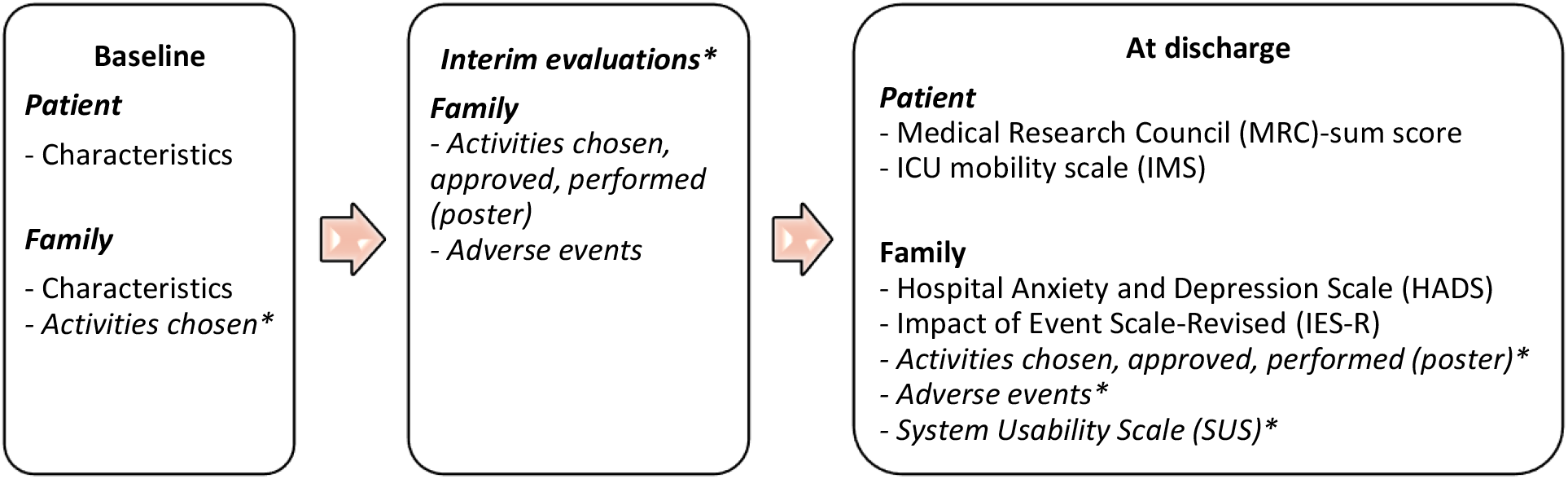
Flowchart of data collection * and in italics: measurements within the intervention group

#### 2.4.2. Additional measurements in intervention group

A number of additional data was collected in the intervention group, shown in figure 1. At baseline, the activities chosen by the family members were recorded. During the interim evaluations and at discharge, data regarding the activities, i.e. activities approved by the physiotherapist and activities performed by family members, were collected from the poster. Additionally, adverse events (e.g. dislocation of endotracheal tubes, lines or other medical equipment, or patient injuries) were assessed by interviewing family members during the interim evaluations, and by copying the events noted on the posters. Last, the System Usability Scale (SUS) was completed by family members at discharge, to assess usability of the brochure and folder. The final SUS score ranges from 0 to 100. Higher scores imply higher usability, and SUS score higher than 68 can be considered above average [29].

### 2.5 Study outcomes

#### 2.5.1 Primary outcome

The primary outcome was feasibility of family participation in physiotherapy-related tasks. The following items were assessed to determine feasibility: 1) recruitment rate; 2) percentage of family members who chose physiotherapy-related tasks at baseline; 3) usage of physiotherapy-related tasks; 4) usability of the poster and brochure; and 5) adverse events. For recruitment rate (1), the percentage of eligible family members who consented to participate in this pilot was calculated, and a percentage of 50% was set a priori as the lower limit. Regarding the percentage family members who chose physiotherapy-related tasks at baseline (2), a percentage of 70% was set as lower limit. Usage of physiotherapy-related tasks (3) was considered as adequate if physiotherapy-related tasks were performed on an average of 70% of all days that it was possible to participate (i.e. the days from start participating to ICU discharge). Additionally, an overview of the executed physiotherapy-related tasks, i.e. the percentages passive, active, functional and breathing exercises, was made. For the usability of the poster and brochure (4) the mean SUS score was analyzed, where a SUS score higher than 68 can be considered above average [29]. Last, no adverse events (5) should be reported by family members. To state that family participation in physiotherapy-related tasks is feasible, we defined a priori that at least four of the five pre-defined criteria had to be met.

#### 2.5.2 Secondary outcomes

As secondary outcomes, two preliminary effectiveness outcomes were compared between the intervention group and control group, at discharge from the ICU. The first outcome was on patient level, i.e. patients’ physical functioning assessed with the MRC-sum score and IMS. As second effectiveness outcome symptoms of anxiety, depression and PTSD were evaluated in family members, using the HADS and IES-R [30-33]. Regarding these two questionnaires, higher scores indicate more symptoms of anxiety, depression and PTSD. In addition to the overall HADS and IES-R scores, the prevalence of family members with symptoms of anxiety or depression, and the prevalence of family members with a post-traumatic stress reaction were examined, where a cutoff HADS score of 8 and greater on the anxiety or depression subscale indicate at least mild anxiety or depression symptoms, and mean IES-R scores of 1.6 and greater indicate a post-traumatic stress reaction with a substantial risk of PTSD [30-33].

### 2.6 Data analysis

All continuous variables were tested for normality with the Kolmogorov-Smirnov test. Participant characteristics were described using descriptive statistics, and tested (intervention group vs. control group) with the chi-square test, Mann Whitney test, or the independent samples t-test, where appropriate. Regarding the primary outcome, descriptive statistics were used to summarize the recruitment rate, usage, usability and adverse events [22]. Regarding the secondary outcomes, possible differences between the intervention group and control group in MRC-sum, IMS, HADS and IES-R scores were tested using the Mann Whitney test or the independent samples t-test, where appropriate. No correction for multiple testing was performed because this was a pilot study. Additionally, differences in the proportion of family members who had a HADS score of 8 or greater in each category, and differences in the percentage of family members with mean IES-R scores of 1.6 and greater between the intervention group and control group were tested, using the chi-square test. As this was a pilot study, the level of missing data was documented but no imputation was undertaken. All data analyses were performed in SPSS.

## 3. Results

### 3.1. Participant characteristics

Between March 20^th^ 2021 and June 21^th^ 2021, 34 family members (19 intervention group, 15 control group) of 27 critically ill patients (16 intervention group, 11 control group) were included. Characteristics of the study population are presented in table 3. The majority of the included family members were partners and female, with a mean age of 49.5 (SD 16.0). Regarding the patients, the majority of the patients were male, with a mean age of 55.3 (SD 14.7). There were no significant differences observed in the characteristics of the family members and patients in the intervention and control group.

**Table 3.** Family and patient demographics.

| Family demographics | Intervention group (n=19) | Control group (n=15) | P-value |
| --- | --- | --- | --- |
| Male, n (%) | 6 (31.6) | 4 (26.7) | 0.755 |
| Age (year), mean (SD) | 48.7 (15.6) | 50.7 (17.1) | 0.734 |
| Relationship to patient, n (%) |  |  | 0.625 |
| - Partner | 11 (57.9) | 9 (60.0) |  |
| - Parent | 2 (10.5) | 0 (0.0) |  |
| - Child | 5 (26.4) | 5 (33.4) |  |
| - Sibling | 1 (5.3) | 1 (6.7) |  |
| Previous ICU experience, n (%) |  |  | 0.409 |
| - Yes | 9 (47.4) | 5 (33.3) |  |
| - No | 10 (52.6) | 10 (66.7) |  |
| Patient demographics | Intervention group (n=16) | Control group (n=11) |  |
| Male, n (%) | 8 (50.0) | 6 (54.5) | 0.816 |
| Age (year), mean (SD) | 51.8 (14.9) | 60.4 (13.4) | 0.137 |
| Reason for ICU admission, n (%) |  |  | 0.288 |
| - Medical | 9 (56.3) | 3 (27.3) |  |
| - Emergency surgery | 6 (37.5) | 6 (54.5) |  |
| - Planned surgery | 1 (6.3) | 2 (18.2) |  |
| Highest SOFA score, mean (SD) <sup>a</sup> | 11.2 (3.0) | 9.4 (1.9) | 0.087 |
| APACHE II score, mean (SD) <sup>b</sup> | 23.9 (6.4) | 18.9 (4.3) | 0.063 |
| Discharge location, n (%) |  |  | 0.242 |
| - General ward | 14 (87.5) | 10 (90.9) |  |
| - Died | 2 (12.5) | 0 (0.0) |  |
| - Rehabilitation center | 0 (0.0) | 1 (9.1) |  |
| Length of stay ICU (days), mean (SD) | 23.3 (14.9) | 19.8 (11.8) | 0.529 |

<sup>a</sup> SOFA score: Sequential Organ Failure Assessment score, score ranges from 0 to 24, and higher scores reflect higher levels of organ dysfunction and a higher risk of death
<sup>b</sup> APACHE II score: Acute Physiology and Chronic Health Evaluation score, score ranges from 0 to 71 and higher scores correspond to more severe disease and a higher risk of death

### 3.2. Feasibility

Regarding recruitment, 75.6% (34 out of 45) of the eligible family members who were asked to participate in this study agreed and were included. The percentage family members in the intervention group who chose physiotherapy-related tasks at baseline, was 94.7% (18 out of 19). Within this group, the total number of days that is was possible to participate in physiotherapy-related tasks ranged from 3 days to 40 days and the number of days that family members actually performed physiotherapy-related tasks ranged from 0 to 18 days. Overall, physiotherapy tasks were executed on an average of 56.7% (SD 30.8) of all days that it was possible to participate. Focusing on the amount and type of activities these family members performed, family members executed physiotherapy-related tasks for 151 times within 15 patients. Of these, 102 times (67.6%) passive movement exercises were done, 42 times (27.8%) it were active limb exercises, 4 times (2.6%) it were functional exercises, and 3 times (2.0%) they executed breathing exercises (figure 2). Regarding usability of the intervention, the mean SUS score was 77.1 (SD 10). No serious adverse events were reported by family members.

**Figure 2.**
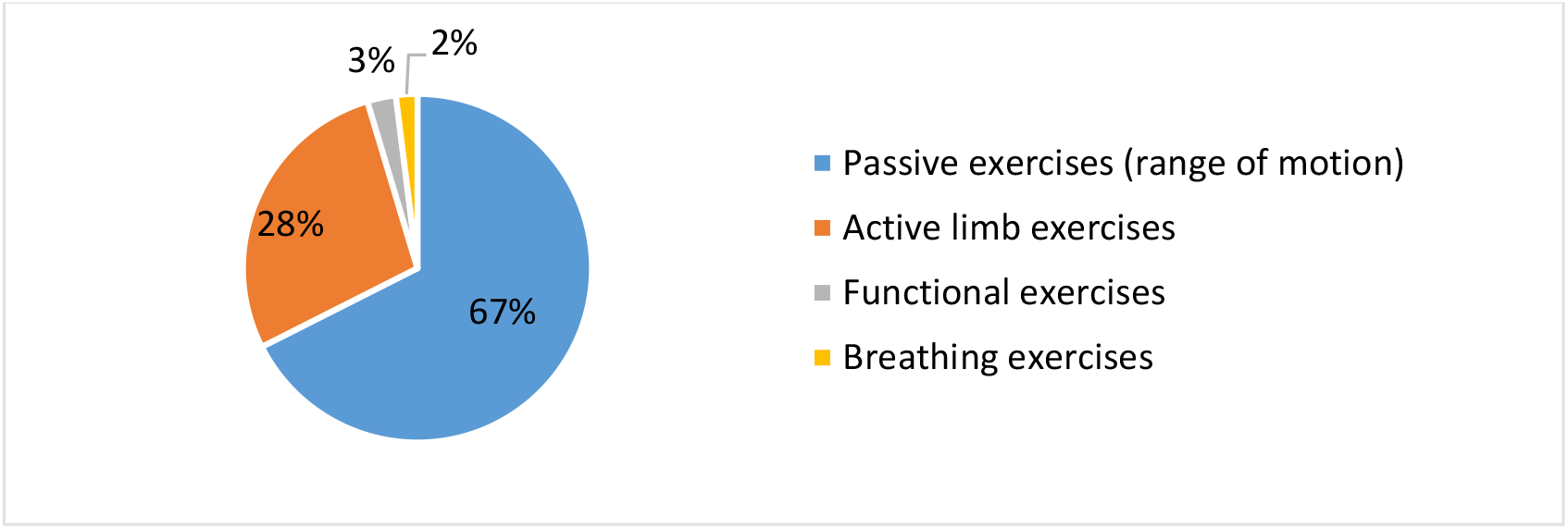
Distribution of physiotherapy-related tasks performed by family members of critically ill patients

### 3.3. Preliminary effectiveness

Family members included in the intervention group had a lower mean HADS-depression score (P=0.030) and mean IES-R score (P=0.047) than family members in the control group (table 4) at ICU discharge. No significant differences were found regarding the other secondary outcomes.

**Table 4.** Differences in patients’ physical functioning and symptoms of anxiety, depression and PTSD, between the intervention group and control group at ICU discharge.

| Outcome measure | Intervention group | Control group | P-value |
| --- | --- | --- | --- |
| MRC-sum score, mean (SD) | 40.0 (10.1) | 37.3 (10.4) | 0.526 |
| IMS, mean (SD) | 3.9 (1.6) | 3.3 (1.6) | 0.345 |
| HADS- anxiety, median (IQR) | 7.0 (5.0-10.0) | 10.0 (5.5-14.0) | 0.215 |
| HADS- depression, mean (SD) | 6.1 (3.3) | 9.2 (4.1) | 0.030* |
| IES-R, mean (SD) | 19.3 (9.7) | 28.0 (13.7) | 0.047* |
| Symptoms of anxiety <sup>a</sup> |  |  | 0.269 |
| - Yes, n (%) | 7 (41.2) | 8 (61.5) |  |
| - No, n (%) | 10 (58.8) | 5 (38.5) |  |
| Symptoms of depression <sup>b</sup> |  |  | 0.153 |
| - Yes, n (%) | 6 (35.3) | 8 (61.5) |  |
| - No, n (%) | 11 (64.7) | 5 (38.5) |  |
| Post-traumatic stress reaction <sup>c</sup> |  |  | 0.197 |
| - Yes, n (%) | 2 (11.8) | 4 (30.8) |  |
| - No, n (%) | 15 (88.2) | 9 (69.2) |  |

<sup>a</sup> total HADS-anxiety score $\geq 8$ , <sup>b</sup> total HADS-depression score $\geq 8$ , <sup>c</sup> mean IES-R score $\geq 1.6$

## 4. Discussion

This pilot study investigated the feasibility of family participation in physiotherapy-related tasks of critically ill patients, and evaluated preliminary effectiveness on patients’ physical functioning and symptoms of anxiety, depression and PTSD in family members. Findings of this study suggest that family participation in physiotherapy-related tasks of critically ill patients is feasible. Additionally, family members in the intervention group reported less symptoms of depression and PTSD.

To determine feasibility, pre-set criteria on recruitment rate, percentage of relatives who chose physiotherapy-related tasks, usage of physiotherapy-related tasks, usability, and adverse events were examined. The results of our pilot demonstrated that four of these five criteria met the predefined requirements, concluding that family participation in physiotherapy-related tasks of critically ill patients seems feasible. Usage of physiotherapy-related tasks did not meet the predefined lower limit. Currently, there are very few comparative studies on the usage and feasibility of participation of family members in physiotherapy-related tasks. One recent study, where nurses completed daily observations, examined how often family participated after implementing an intervention including a menu with activities for family involvement in hands-on ICU care. Their observations showed that family executed touching activities (e.g. massage, applying lotion) on average 64% of all days, and personal care activities (e.g. assist with turning and positioning) on average 45% of all days, which is in line with our percentage for usage [34]. Possibly it is not feasible for family to participate on a daily basis, however, reasons for this must be further investigated. Additionally, our results showed that family members mainly executed passive movement exercises. To increase the impact of patients’ physical functioning by family participation, it is important family members participate frequently and in active exercises. A reason why active exercises were executed less than passive exercises could be that many critically ill patients are sedated in the first days of the ICU admission [35], resulting in more possibilities for passive movements than active exercises. Afterwards, critically ill patients are often delirious. Delirium occurs in up to 50% of ICU patients during their admission [36, 37], which can make it hard for family members to execute active limb exercises. Additionally, it might be that family members need more personal guidance and training during the time, since it has been demonstrated that their needs and behavior regarding participating in promoting patients’ physical activity change during the ICU admission from passive to more proactive [19][38]. Unfortunately, we did not examine these factors of influence on the usage and thereby feasibility of active limb exercises in this study.

Focusing on the preliminary effectiveness on patient level, our study did not find significant differences in patients’ physical functioning at discharge. This may be due to the small sample size and fact that physiotherapy-related tasks, which were mostly passive movements, were performed about half of all possible days. Involving family members in interventions such as active limb exercises, and thereby increase the frequency of physical activity during the day, may promote patients’ physical recovery [19]. Since physical rehabilitation starts early in the ICU it is best to start with this intervention there, but to have more impact on patients’ physical functioning we suggest to continue the intervention after discharge from the ICU, at general wards, where physical rehabilitation continues and family participation in active exercises is possibly more feasible since factors as sedation and delirium play less of a role.

Regarding the preliminary effectiveness on family members, symptoms of depression and PTSD at ICU discharge were lower in the intervention group compared to the control group. Symptoms of anxiety, the prevalence of family members with at least mild symptoms of anxiety or depression, and the prevalence of family members with a post-traumatic stress reaction did not show significant differences. However, these outcomes on family level may be influenced by the design (non-equivalent control group with small sample size, assessment of stress-related symptoms solely at ICU discharge) and/or baseline characteristics. For example, the intervention group included more family members with previous experience in the ICU, these family members might have less stress-related symptoms at baseline, resulting in even fewer symptoms at discharge. Currently, only a few studies have evaluated interventions for family members to be involved in the care of adult ICU patients with the goal of reducing stress-related symptoms in family members [34, 39, 40]. These studies investigated outcomes related to long-term psychological distress, where we examined short-term stress-related symptoms, and mostly reported prevalence percentages with variating effects on symptoms of anxiety, depression, and PTSD [34, 39, 40]. To our knowledge, there are no studies who have evaluated an intervention for family participation in the ICU assessing stress-related symptoms at ICU discharge, which makes it hard to compare our results. Since it is proven that symptoms of anxiety and depression in family members of critically ill patients decrease over time, and disorders as an anxiety/depression disorder and PTSD often arises at a later stage, is recommended to assess anxiety, depression and PTSD-related symptoms in family members from two months after ICU discharge [9, 41, 42]. Therefore, we suggest future studies investigating the effectiveness of interventions for family participation in physiotherapy-related care to evaluate long-term effects on the prevalence and symptoms of PTSD, anxiety and depression [42].

A major strength of this study is the design with only few exclusion criteria, which suggests the results may apply to family members of a wide variety of patients admitted to an adult mixed ICU. There are, however, also several limitations in the design that may have impacted the results. First, we conducted a quasi-experimental study with a non-equivalent control group design. There likely were unmeasured factors between the two groups that limit the interpretation of the results, and external factors that may have influenced the outcomes could not be ruled out. In addition, the non-equivalent control group design, whereby family members were allowed to choose to participate or not, may have resulted in selection bias. The control group experienced more symptoms of depression and PTSD, which may be because family members with more stress-related symptoms did not want to participate in activities and therefore entered the control group. However, family members were mostly enthusiastic and wanted to participate in activities, which accelerated the inclusion of the intervention group and the last 12 family members were only included in the control group, resulting in less selection bias. Nevertheless, to avoid this limitation, larger sample sizes are recommended in future studies, studies must contain a proper control group, and should also assess stress-related symptoms at baseline. Second, the researcher (LvD) who approached the participants also worked as physiotherapists in the ICU, resulting in the possibility that there was a prior relationship to the patient and/or family as part of clinical care. Future large scale clinical trials on this topic should be conducted by an independent researcher. Besides, we did not identify the possible reasons why family members did not participate in physiotherapy-related tasks on a daily basis. One of these reasons may be that family members did not visit their loved one every day, or that family members needed more personal guidance during the time, or that the patient did not appreciate to exercise during family visits. Additional research, using interviews or surveys, is needed to identify these factors influencing the usage and thereby feasibility of family participation in physiotherapy-related tasks before adjusting the intervention and performing a large scale clinical trial. Third limitation to mention is that this study is conducted during the COVID-19 pandemic. Due to the pandemic, elements of the ICU care were momentary changed. For example, family members of isolated COVID-19 patients were not allowed to visit the ICU daily, and therefore excluded for the intervention and pilot study. Since many ICU beds were used by COVID-19 patients during the pilot and many surgeries had to be postponed [43], the enrollment was likely lower than if there were no COVID-19 patients in the ICU. In addition, the visitation of loved ones in the regular ICU was temporary minimized to a maximum of two different persons per 24 hours, which might have resulted in less possibilities for family involvement. These factors together may negatively have influenced the pilot study and affected the feasibility of family participation during this time period. Probably, family participation in the ICU will be more applicable in the post-pandemic world than during this study period.

In conclusion, the results of this pilot study suggest that family participation in physiotherapy-related tasks of critically ill patients is feasible. Since this is the first study focusing on this specific topic, more research is needed to thoroughly evaluate the effectiveness of an intervention for family participation in physiotherapy-related tasks on patients’ physical functioning and long-term symptoms of anxiety, depression and PTSD in family members.

## Clinical Messages

- It seems feasible to involve family members of critically ill patients in performing physiotherapy-related tasks
- An user-centered intervention is needed to increase family participation in physiotherapy-related tasks

## Data Availability

All data produced in the present study are available upon reasonable request to the authors

## Acknowledgements

Not applicable

## Consent for publication

Not applicable

## Ethical approval and consent to participate

The study protocol was assessed and approved by the medical ethics committee of the UMC Utrecht (study protocol number 21-071)

## References

1. Hermans G and Van den Berghe G, Clinical review: intensive care unit acquired weakness. Critical Care, 2015. 19(1): p. 274.

2. Monika C. Kerckhoffs, I.W.S., Annemiek E. Wolters, Lotte Kok, Marike van der Schaaf en Diederik van Dijk, Langetermijnuitkomsten van IC-behandeling. Nederlands tijdschrift voor geneeskunde, 2016.

3. Judy E. Davidson, D., RN, Fccm, CNS; Maurene A. Harvey Mph, RN, MCCM; Jessica Schuller, Bsn, RN; and Gary Black, Med, BSEd, Bfa, AA, Post-intensive care syndrome: What it is and how to help prevent it. American Nurse Today. 8(5).

4. Van der Schaaf M. B.A., Dongelmans DA, Vroom MB, Nollet F., Functional status after intensive care: a challenge for rehabilitation professionals to improve outcome. J Rehabil Med., 2009. 41.

5. Herridge MS, T.C., Matté A, Tomlinson G, Diaz-Granados N, Cooper A, Guest CB, Mazer CD, Mehta S, Stewart TE, Kudlow P, Cook D, Slutsky AS, Cheung AM; Canadian Critical Care Trials Group., Functional disability 5 years after acute respiratory distress syndrome.. N Engl J Med., 2011. 364(12): p. 1293–304.

6. Herridge MS. C.A., Tansey CM, Matte-Martyn A, Diaz-Granados N, Al-Saidi F, Cooper AB, Guest CB, Mazer CD, Mehta S, Stewart TE, Barr A, Cook D, Slutsky AS; Canadian Critical Care Trials Group., One-year outcomes in survivors of the acute respiratory distress syndrome. N Engl J Med, 2003. 348(8): p. 683–93.

7. Needham, D.M., et al., Improving long-term outcomes after discharge from intensive care unit: report from a stakeholders’ conference. Critical Care Medicine, 2012. 40(2): p. 502–509.

8. Alfheim HB, et al., Post-traumatic stress symptoms in family caregivers of intensive care unit patients: A longitudinal study. Intensive Crit Care Nurs, 2019. 50: p. 5–10.

9. Pochard F, et al., Symptoms of anxiety and depression in family members of intensive care unit patients: ethical hypothesis regarding decision-making capacity.. Critical Care Medicine, 2001. 29(10).

10. McAdam JL, et al., Psychological Symptoms of Family Members of High-Risk Intensive Care Unit Patients. Am J Crit Care 2012. 21(6): p. 386–394.

11. Needham DM, et al., Early physical medicine and rehabilitation for patients with acute respiratory failure: a quality improvement project. Arch Phys Med Rehabil, 2010. 91(4): p. 536–42.

12. Parker A, Sricharoenchai T, and Needham DM, Early Rehabilitation in the Intensive Care Unit: Preventing Physical and Mental Health Impairments. Curr Phys Med Rehabil Rep, 2013. 1(4): p. 307–314.

13. Sommers J. E.R., Dettling-Ihnenfeldt D, et al., Physiotherapy in the intensive care unit: an evidence-based, expert driven, practical statement and rehabilitation recommendations.. Clin Rehabil, 2015. 29: p. 1051–1063.

14. Kayambu G. B.R., Paratz J., Early physical rehabilitation in intensive care patients with sepsis syndromes: a pilot randomised controlled trial.. Intensive Care Med, 2015. 41: p. 865–874.

15. D., N., Mobilizing Patients in the Intensive Care Unit Improving Neuromuscular Weakness and Physical Function.. JAMA, 2008. 300: p. 1685–1690.

16. Olding M, et al., Patient and family involvement in adult critical and intensive care settings: a scoping review. Health Expect, 2016. 19(6): p. 1183–1202.

17. Haines KJ, Engaging Families in Rehabilitation of People Who Are Critically Ill: An Underutilized Resource. Physical Therapy, 2018. 98(9): p. 737–744.

18. Mackie, B.R., M. Mitchell, and P.A. Marshall, The impact of interventions that promote family involvement in care on adult acute-care wards: An integrative review. Collegian, 2018. 25(1): p. 131–140.

19. Lmm, v.D., Qualitative study-under review Physiother Theory Pract., 2021.

20. van Delft LMM, et al., Family participation in physiotherapy-related tasks of critically ill patients: A mixed methods systematic review.. J Crit Care., 2021. 62: p. 49–57.

21. Craig P. D.P., Macintyre S, Michie S, Nazareth I, Petticrew M,, Developing and evaluating complex interventions: the new Medical Research Council guidance. BMJ: British Medical Journal 2008. 337: p. a1655.

22. Lancaster, G.A., S. Dodd, and P.R. Williamson, Design and analysis of pilot studies: recommendations for good practice. Journal of Evaluation in Clinical Practice, 2004. 10(2): p. 307–312.

23. Hermans G. C.B., Vanhullebusch T, Segers J, Vanpee G, Robbeets C, et al., Interobserver agreement of Medical Research Council sum-score and handgrip strength in the intensive care unit. Muscle Nerve., 2012. 45(1): p. 18–25.

24. AS, Z. and S. RP, The hospital anxiety and depression scale Acta Psychiatrica Scandinavica, 1983. 67: p. 361–370.

25. J.M. Turner-Cobb, P.C.S. P. Ramchandani, F.M. Begen & A. Padkin, The acute psychobiological impact of the intensive care experience on relatives,. Psychology, Health & Medicine, 2016. 21(1): p. 20–26.

26. Beck JG. G.D., Read JP, Clapp JD, Coffey SF, Miller LM, Palyo SA., The impact of event scale-revised: psychometric properties in a sample of motor vehicle accident survivors.. J Anxiety Disord., 2008. 22(2): p. 187–98.

27. Weiss, D.M.. CR., The impact of event scale-revised. Assessing Psychological Trauma and PTSD., 1997: p. 399–411.

28. Creamer M. B.R., Failla S., Psychometric properties of the Impact of Event Scale-Revised.. Behav Res Ther., 2003. 41: p. 1489–1496.

29. JR., L., The system usability scale: past, present, and future. Int J Hum Comput Interact. 2018. 34: p. 577–90.

30. Stevenson JE. C.E., Bienvenu OJ, et al., General anxiety symptoms after acute lung injury: predictors and correlates.. J Psychosom Res, 2013. 75(3): p. 287–293.

31. Bienvenu OJ. W.J., Yang A, Hopkins RO, Needham DM., Posttraumatic stress disorder in survivors of acute lung injury: evaluating the Impact of Event Scale-Revised.. Chest, 2013. 144(1): p. 24–31.

32. Needham DM. S.K., Dinglas VD, Chessare CM, Friedman LA, Bingham CO 3rd, Turnbull AE., Core Outcome Measures for Clinical Research in Acute Respiratory Failure Survivors. An International Modified Delphi Consensus Study.. Am J Respir Crit Care Med., 2017. 196(9): p. 1122–1130

33. Chan KS. A.F.L., Bienvenu OJ, Dinglas VD, Cuthbertson BH, Porter R, Jones C, Hopkins RO, Needham DM., Distribution-based estimates of minimal important difference for hospital anxiety and depression scale and impact of event scale-revised in survivors of acute respiratory failure Gen Hosp Psychiatry, 2016. 42(32-5).

34. Amass, T.H., et al., Family Care Rituals in the ICU to Reduce Symptoms of Post-Traumatic Stress Disorder in Family Members-A Multicenter, Multinational, Before-and-After Intervention Trial. Critical Care Medicine, 2020. 48(2): p. 176–184.

35. Reade MC. F.S., Sedation and delirium in the intensive care unit. N Engl J Med., 2014. 370(5): p. 444–54.

36. Wolters AE. S.A., van der Kooi AW, van Dijk D., Cognitive impairment after intensive care unit admission: a systematic review. Intensive Care Med 2013. 39: p. 376–386.

37. Zaal IJ. S.A., Delirium in critically ill patients: epidemiology, pathophysiology, diagnosis and management. Drugs 2012, 2012. 72: p. 1457–1471.

38. Felten-Barentsz KM, v.d.W.-v.D.V., Vloet L, Koenders N, Nijhuis-van der Sanden MWG, Hoogeboom TJ., Family participation during physical activity in the intensive care unit: A longitudinal qualitative study. J Crit Care, 2021. 65: p. 42–48.

39. Mitchell ML. C.F., Kean S, Stone R, Murfield J, Dwan T., Patient, family-centred care interventions within the adult ICU setting: An integrative review.. Aust Crit Care, 2016. 29(4): p. 179–193

40. White, D.B., et al., A Randomized Trial of a Family-Support Intervention in Intensive Care Units. New England Journal of Medicine, 2018. 378(25): p. 2365–2375.

41. Azoulay E. P.F., Kentish-Barnes N, Chevret S, Aboab J, Adrie C, et al. FAMIREA Study Group., Risk of post-traumatic stress symptoms in family members of intensive care unit patients Am J Respir Crit Care Med., 2005. 171(9): p. 987–94.

42. Davidson JE, Jones C, and Bienvenu OJ, Family response to critical illness: postintensive care syndrome-family. Critical Care Medicine, 2012. 40(2): p. 618–24.

43. SKR. SKR impact report 2021.

